# Safety, tolerability, and immunogenicity of a new SARS-CoV-2 recombinant Gamma variant RBD-based protein adjuvanted vaccine, used as heterologous booster in healthy adults: a Phase 1 interim report

**DOI:** 10.1101/2023.01.10.22284009

**Authors:** Karina A. Pasquevich, Lorena M. Coria, Ana Ceballos, Bianca Mazzitelli, Juan Manuel Rodriguez, Agostina Demaría, Celeste Pueblas Castro, Laura Bruno, Lucas Saposnik, Melina Salvatori, Augusto Varese, Soledad González, Veronica V. González Martínez, Jorge Geffner, Diego Álvarez, Laboratorio Pablo Cassará R&D and CMC group for ARVAC-CG, Ethel Feleder, Karina Halabe, Pablo E. Perez Lera, Federico Montes de Oca, Julio C. Vega, Mónica Lombardo, Gustavo A. Yerino, Juan Fló, Juliana Cassataro

**Affiliations:** Instituto de Investigaciones Biotecnológicas, Universidad Nacional de San Martín (UNSAM) – Consejo Nacional de Investigaciones Científicas y Técnicas (CONICET). San Martín (1650), Buenos Aires, Argentina; Escuela de Bio y Nanotecnologías (EByN), Universidad Nacional de San Martín. San Martín (1650), Buenos Aires, Argentina; Instituto de Investigaciones Biomédicas en Retrovirus y SIDA, INBIRS-CONICET, Facultad de Medicina UBA, Buenos Aires, Argentina; Fundación Pablo Cassará - Unidad de I+D de Biofármacos. Saladillo 2452 C1440FFX, Ciudad Autónoma de Buenos Aires, Argentina; Ministerio de Salud de la Provincia de Buenos Aires, Buenos Aires, Argentina; Laboratorio Pablo Cassará – Unidad de I+D de Biofármacos. Saladillo 2452 C1440FFX, Ciudad Autónoma de Buenos Aires, Argentina; FP CLINICAL PHARMA, Ciudad Autónoma de Buenos Aires, Argentina; Nobeltri. Ciudad Autónoma de Buenos Aires, Argentina

**Keywords:** COVID-19 subunit vaccine, Gamma variant, Phase 1, SARS-CoV-2, heterologous booster

## Abstract

**Background:** In view of the emergence of SARS-CoV-2 immune escape variants and evidence of waning immunity, new immunisation strategies and variant-adapted vaccines are needed. Based on preclinical proof of concept studies and requirement of variant-adapted and booster vaccines, the Gamma Variant RBD-based ARVAC-CG vaccine was selected for a first clinical trial in humans.

**Methods:** Eighty participants (healthy adults, 18-55 years-old) were sequentially assigned to receive two (28 days apart) intramuscular doses of 25-μg (n=60) or 50-μg (n=20) of a Gamma RBD-based subunit vaccine adjuvanted with aluminium hydroxide. The primary endpoint was safety. The secondary objective was to describe the neutralising antibody response against the SARS-CoV-2 Ancestral strain and several variants of concern (Gamma, Delta, Omicron BA.1 and Omicron BA.5) measured by a live virus-based neutralisation assay. Cellular immune responses were studied as an exploratory objective by an enzyme-linked immunospot (ELISpot) assay. This trial is registered in ClinicalTrials.gov (<u>NCT05656508</u>).

**Findings:** The interim results from the ongoing phase 1 study are described. ARVAC-CG exhibited a satisfactory safety profile, a robust and broad booster response of neutralising antibodies against the Ancestral strain of SARS-CoV-2, the Gamma variant, and other VOCs (Delta, Omicron BA.1 and Omicron BA.5) and a booster effect on T cell immunity.

**Interpretation:** ARVAC-CG is safe and highly immunogenic when used as booster in individuals previously immunised with different COVID-19 vaccine platforms. These results warrant further clinical evaluation of this vaccine candidate for boosting other COVID-19 vaccines.

**Funding:** Laboratorio Pablo Cassará S.R.L. (Argentina).

**Research in context:** *Evidence before this study:* Next-generation COVID-19 vaccines are based on a variant-adapted approach, using a strain other than the parental strain of SARS-CoV-2 (Wuhan or D614G strain). It has been suggested that the use of vaccines containing Beta spike protein may be an interesting strategy to acquire wider protection against SARS-CoV-2 variants. The Beta variant has been tested as booster in different monovalent or bivalent vaccine platforms. Indeed, Sanofi and GSK VidPrevtyn® Beta has recently been approved in Europe representing the first protein-based next-generation COVID-19 booster vaccine. While the receptor binding domain (RBD) of the spike protein of Gamma and Beta SARS-CoV-2 variants are very similar, no clinical data on Gamma variant-based COVID-19 vaccines has been published so far. Preclinical data in mice indicate that the Gamma variant-based vaccine is more immunogenic and induces a broader nAb response than the ancestral RBD-based vaccine.

*Added value of this study:* To our knowledge, these is the first clinical trial reported from any monovalent Gamma variant RBD protein adjuvanted vaccine used as heterologous booster of different primary series vaccine platforms. Two different vaccine doses were tested, and both exhibited a good profile of safety, tolerability and reactogenicity. ARVAC-CG as a single heterologous booster induced a significant increase of broad-spectrum neutralising antibodies against Ancestral, Gamma, Delta, Omicron BA.1 and Omicron BA.5 variants of concern (VOCs), binding antibodies, and IFN-γ producing cells. All these immune responses were significantly boosted in individuals primed with vaccines from different platforms. Plasma from vaccinees receiving a heterologous booster with ARVAC-CG was superior to plasma from BTN16b2 boosted individuals in neutralising Omicron BA.1 and BA.5 SARS-CoV-2 VOCs.

*Implications of all the available evidence:* Here, we present the available data from the phase I study of ARVAC-CG vaccine, involving healthy adults who had previously received a complete primary vaccination schedule with a COVID-19 vaccine. The positive safety and immunogenicity results of the ARVAC-CG vaccine candidate presented here justify further evaluation of its immunogenicity against currently circulating SARS-CoV-2 VOCs in a comprehensive Phase 2/3 trial. Further research is required to assess the antibody persistence over time after a booster dose of ARVAC-CG.

## INTRODUCTION

The severe acute respiratory virus 2 (SARS-CoV-2) was first identified in November 2020 and soon thereafter, emerging new viral variants dramatically impacted the dynamics of Coronavirus disease 2019 (COVID-19) spread, globally. These virus variants are, in general, more contagious, and virulent than previous strains. Moreover, some variants are capable of immunological and/or therapeutic escape.^1,2^

Numerous vaccines have been developed and proved effective to protect against severe disease, hospitalization, and fatal outcomes.^3-5^ In Argentina, several platforms of COVID-19 vaccines have been introduced, including inactivated vaccines (BBIBP-CorV), viral vectored vaccines (Sputnik V, AZD1222, CanSino), and mRNA vaccines (BNT162b2 and mRNA-1273), resulting in a significant coverage of the population with complete primary vaccine series.^6,7^ However, due to waning immunity and emergence of highly transmissible immune escape viral variants, two-dose COVID-19 vaccination programs may have not been enough to prevent breakthrough infections caused by these variants.^8,9^ Clinical studies suggest that boosting with variant-adapted vaccines would optimise vaccine efficacy (VE) inducing strong and broad immune responses.^10-12^

The pandemic has disproportionately affected low- and middle-income countries (LMICs), which make up about 85% of the world population. Therefore, pandemic remains threat unless most people get vaccinated. In response to the constraints imposed by the COVID-19 pandemic, and the limited access of many Latin American countries to costly vaccines, Laboratorio Pablo Cassará S.R.L., an Argentinian pharmaceutical company, launched a vaccine development program against SARS-CoV-2. ARVAC-CG is an receptor binding domain (RBD) based protein aluminium hydroxide adjuvanted vaccine candidate that was designed and produced entirely in Argentina to be used as booster or primary vaccine against SARS-CoV-2. The vaccine is a variant-adapted vaccine based on the highly immune evasive Gamma SARS-CoV-2 variant of concern (VOC). Non-clinical studies of this vaccine prototype in mice indicated that the Gamma-variant vaccine-candidate is more immunogenic and induces a broader nAb response than the Ancestral vaccine-candidate.^13^

In this interim report safety and immunogenicity data after a booster dose of ARVAC-CG vaccine from an ongoing first in human phase 1 study are presented.

## METHODS

### Trial design and oversight

This trial is registered in ClinicalTrials.gov (<u>NCT05656508</u>). The trial was conducted at Clinical Pharma (Clínica CIAREC, Intense Life S.A, Buenos Aires, Argentina). In this open-label, first-in-human, dose-escalation, phase 1 clinical trial, eligible volunteers were healthy men and nonpregnant women, aged 18 to 55, with a body-mass index of 18 to 30. Health status, assessed during the screening period, was based on medical history and extensive clinical laboratory tests, vital signs, and physical examination. Participants with a history of SARS-Cov-2 infection or COVID-19 within 60 days prior to recruitment into the study, or who tested positive in real-time polymerase-chain-reaction (RT-PCR) assay screening or worked in an occupation with high risk of exposure to SARS-CoV-2, as well as those with an incomplete COVID-19 vaccine primary schedule or who had received a booster dose of any COVID-19 vaccine, were excluded. All participants provided written informed consent before enrolment in the trial. The trial protocol was approved on March 09, 2022, by the Ethic Committee in Clinical Research of Centro de Estudios Infectológicos S.A., and by the Food and Drugs National Regulatory Agency (Administración Nacional de Medicamentos, Alimentos y Tecnología Médica, ANMAT), on March 22, 2022.

The study protocol was initiated on April 10, 2022. Twenty seven out of 107 volunteers (13 males, 14 females) were excluded for not complying with eligibility criteria. Participants were recruited between April 28 and June 23, 2022, and sequentially assigned to one of two vaccine groups, one receiving a 25-μg dose of ARVAC-CG (Group A) and the other a 50-μg dose (Group B). Of the 80 volunteers who received at least one intramuscular dose of ARVAC-CG in the deltoid, three were excluded 28 days after the first dose of the vaccine due to impossibility to complete the protocol for personal reasons. Of the 77 remaining, 59 were inoculated with two 25-μg vaccine doses, and 18 with two 50-μg vaccine doses. Investigators, and laboratory personnel involved in assays were blind to assignment until the end of the follow-up period (Figure 1).

**Figure 1.**
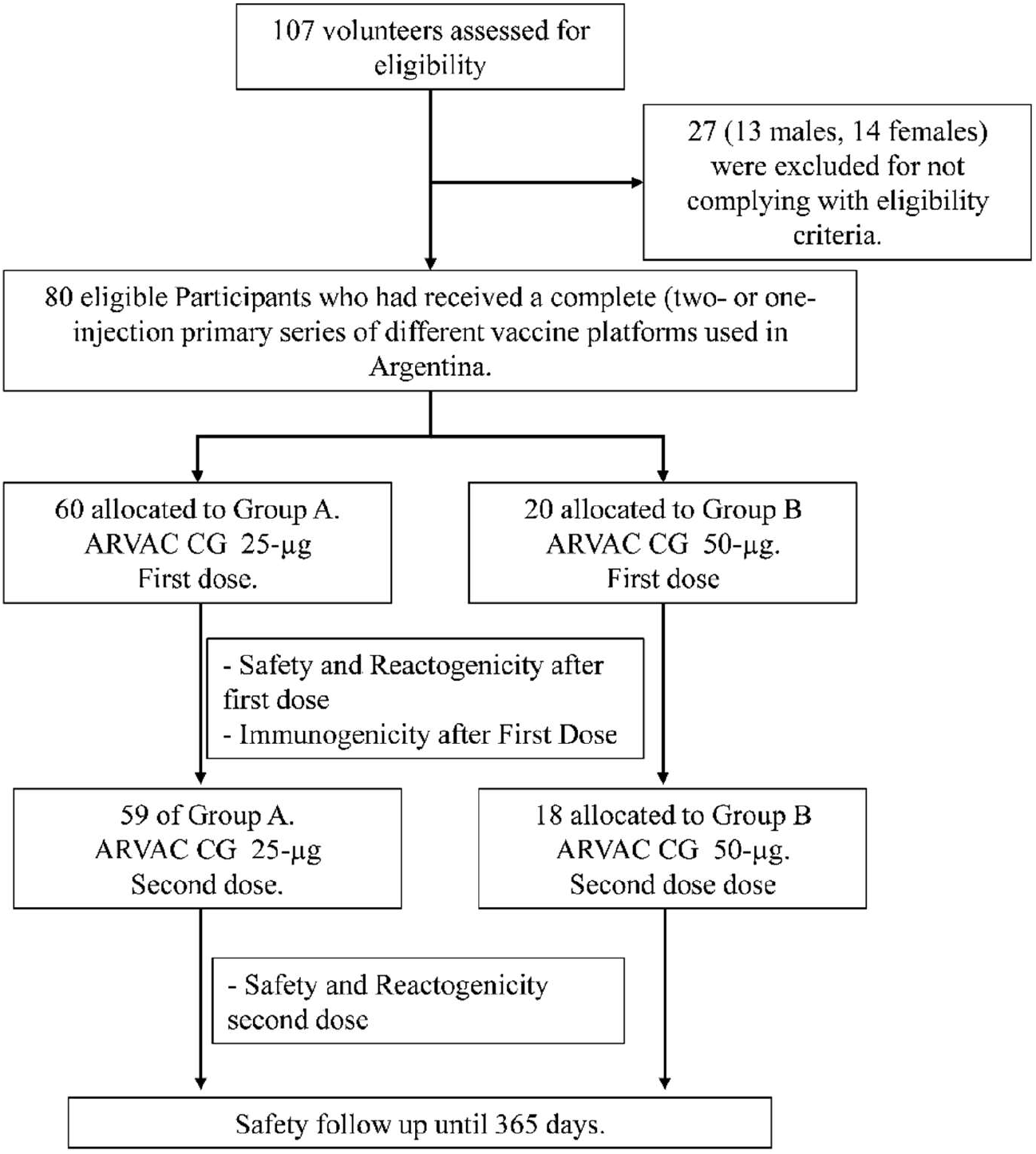
Subject disposition in Phase-1 study (consort diagram). Trial profile showing the study groups, number of participants, and vaccine dose received.

Demographic characteristics of the participants are presented in Table 1. Missing data or deviations from original protocol were infrequent and inconsequential.

**Table 1.**
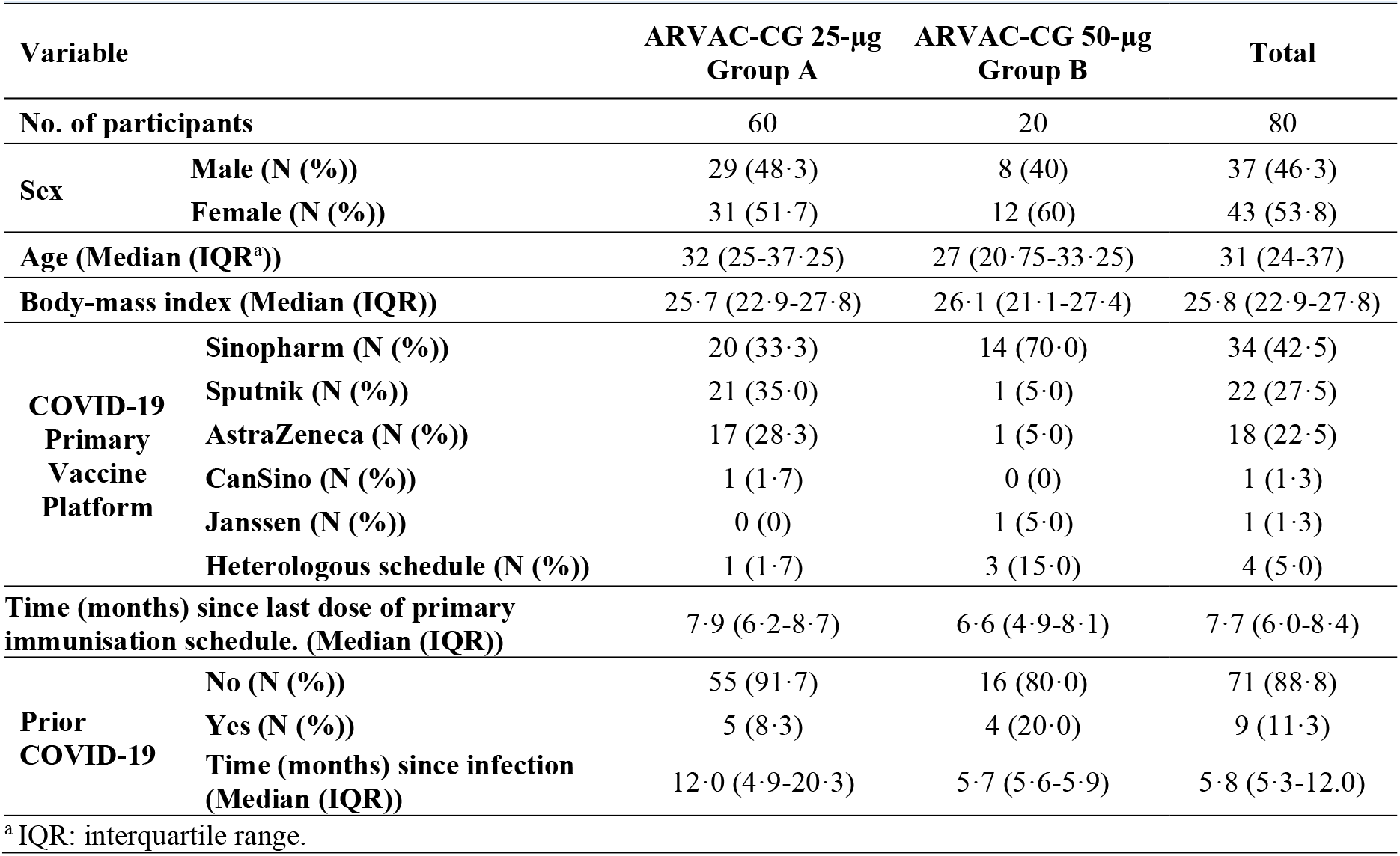
Demographic Characteristics of the Participants in the ARVAC-F1-001 Trial at Enrolment.

Safety was assessed according to the scheme established in the Guidance for Industry, Toxicity Grading Scale for Healthy Adult and Adolescent Volunteers Enrolled in Preventive Vaccine Clinical Trials.^14^ Solicited local and systemic adverse events (AE) were recorded during the first 7 days after each dose of vaccine received, unsolicited events were recorded during the first 28 days after vaccination, laboratory tests were carried out after 7 and 28 days of each dose. The following symptom grading was used for local and systemic AE: grade 1 (mild) to grade 4 (potentially life-threatening). All safety information collected was available to an external Independent Committee of Data Review for continuous monitoring of any relevant AE and recommendations of modifying or interrupting the protocol as necessary.

Causality assessment of AE was based on the standard definition and application of terms for vaccine pharmacovigilance as stated by the Report of the CIOMS/WHO Working Group on Vaccine Pharmacovigilance^15^.

The immunogenicity of ARAVAC CG was compared to 18 samples obtained during the COVID-19 serology surveillance strategy implemented by the Ministry of Health of the Province of Buenos Aires, between 4 February and 31 March 2022. These were individuals with similar demographic characteristics who received a heterologous booster with the Ancestral-based BNT16b2 mRNA vaccine administered at least 3 months after a two-dose primary schedule (Appendix Table S1).

### SARS-CoV-2 Neutralisation assays

Neutralisation assays were performed using live SARS-CoV-2 virus isolates. Serial dilutions of plasma samples from 1/8 to 1/16384 were incubated 1 h at 37°C in the presence of Ancestral (B.1), Gamma, Delta, or Omicron variants (BA.1 or BA.5) in Dulbecco’s Modified Eagle Medium (DMEM), 2% foetal bovine serum (FBS). Then, 50μL of the mixture were deposited over Vero cell monolayers for an hour at 37°C (MOI, 0.01). Infectious medium was removed and replaced by DMEM, 2%-FBS. After 72 h, cells were fixed with 4% paraformaldehyde (4°C, 20 min) and stained with crystal violet solution in methanol. The cytopathic effect (CPE) on the cell monolayer was assessed visually. If damage to the monolayer was observed in the well, it was considered manifestation of CPE and the neutralisation titre was defined as the highest serum dilution that prevented any CPE.

The nAb titres were transformed to international units per ml (IU/ml) by the inclusion in each plate of a secondary standard that was calibrated with the WHO international standard (NIBSC code: 20/268) following the WHO procedures manual.^16^

### SARS-CoV-2 antibody ELISA

Antibodies against SARS-CoV-2 spike protein or RBD were detected using established, commercially available, two-step ELISAs COVIDAR,^14^ or SARS-CoV-2 (RBD) total Ab ELISA from DRG International (DRG Inc, Springfield, NJ 07081 USA). The immunoglobulin G (IgG) concentration of each sample, expressed in Binding Antibody Units/mL (BAU/mL) was calculated by extrapolation of the optical density at 450 nm (OD450) on a calibration curve built using serial dilutions of the WHO International Standard for anti-SARS-CoV-2 immunoglobulin.

### ELISpot

The T-cell mediated immune response against the SARS-CoV-2 RBD was assessed after the *in vitro* peptide stimulation of peripheral blood mononuclear cells (PBMC), followed by IFN-γ and IL-4 enzyme-linked immune absorbent spot (IFN-γ and IL-4 ELISpot, BD Biosciences). A peptide pool of overlapping SARS-CoV-2 peptides, encompassing the SARS-CoV-2 spike RBD covering the Gamma variant was used in the assay (JPT Peptide Technologies GmbH, Germany).

### Statistical analysis

Variables are reported as absolute numbers, percentages, means, medians, and interquartile range (IQR), or geometric means and 95% confidence intervals (CI). Nonparametric statistical tests were used to analyse differences in categorical variables. Immunogenicity response was assessed by means of nAb geometric mean titres (GMTs), and seroconversion rates. Seroconversion was defined as a change in value from below the lower limit of detection (LLOD) to at least 4 or 10 times above, or an increase by a factor of at least four or ten if the baseline value was greater than or equal to the LLOD in comparison to the pre-vaccination baseline value. Differences in mean, geometric mean, or percentage values between groups and among prior vaccine platforms were assessed by means of Mann Whitney u test, Wilcoxon pair-matched test, Kruskal-Wallis test with post hoc Dunn’s for multiple comparisons, Fisher exact test, or Chi square distribution, as appropriate. 95% CI were calculated using the Wilson/Brown method. No assumptions were made for missing data. Statistical analyses were done using GraphPad Prism v8.4.2 (GraphPad Software, San Diego, CA), Two-sided P values <0·05 were considered statistically significant.

## RESULTS

### Population characteristics and local and systemic adverse events

The flowchart of the study is shown in figure 1. Demographic characteristics of the participants are presented in table 1. Vaccination with ARVAC-CG was well tolerated, with mild-to-moderate reactogenicity profiles (Figure 2A). Overall, solicited local AE were more frequently noticed after the first dose of the vaccine than after the second. At least one local AE was observed in 68·3% of Group A volunteers (25-μg vaccine) after the first administration and in 47·5% following the second (p=0·026). In group B (50-μg vaccine), 60·0% of volunteers reported at least one local AE after the first injection whereas 27·8% following the second (p=0·0585) (Table 2).

**Figure 2.**
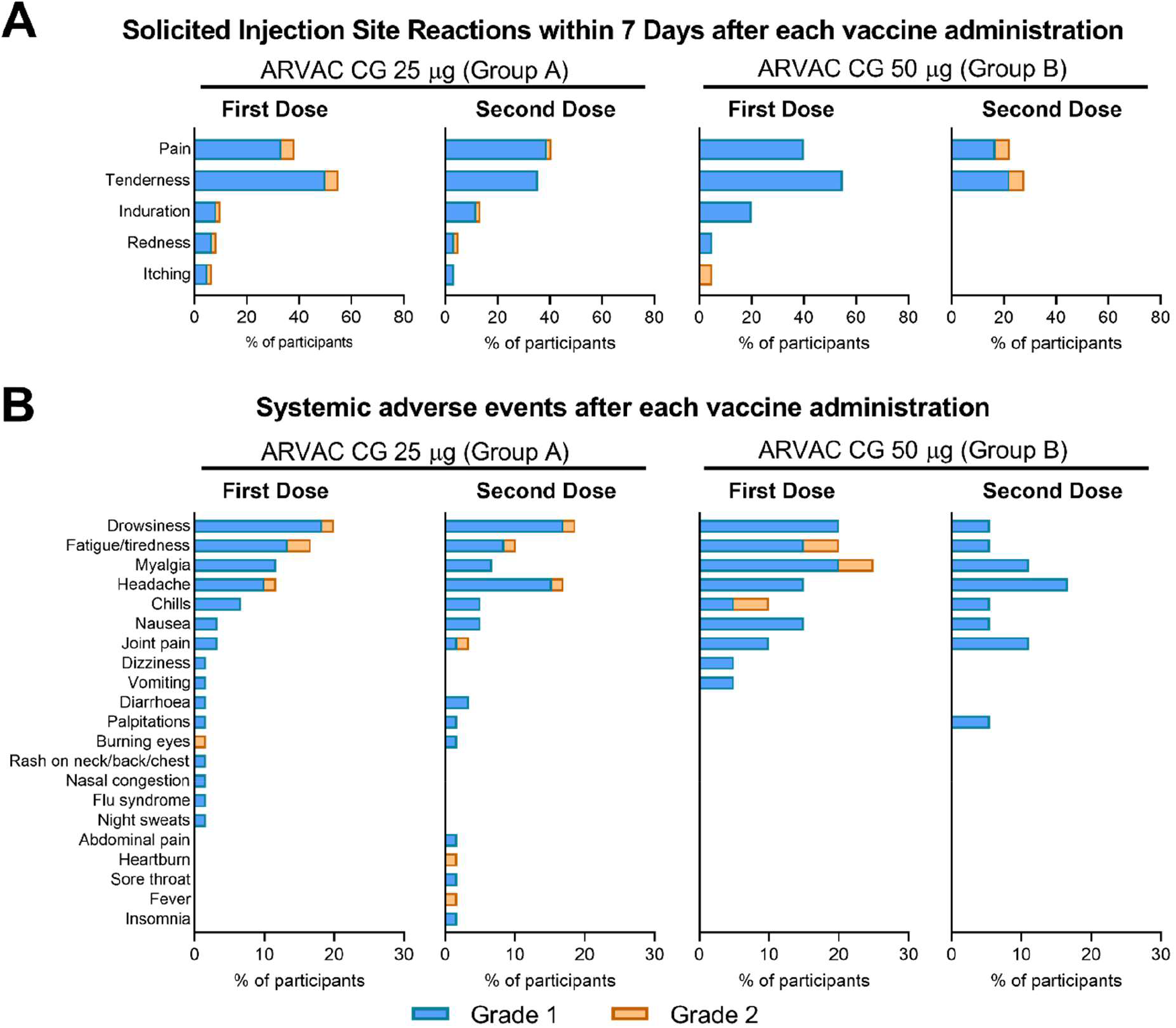
Safety profile. **(A)** Percentage of participants in each study group with the indicated injection site AEs up to 7 days after the first or the second injection. **(B)** Percentage of participants in each study group with the indicated systemic AEs recorded up to 28 days after each vaccine administration. Events were classified according to the FDA toxicity grading scale for healthy adult and adolescent volunteers enrolled in preventive vaccine clinical trials (Mild (Grade 1), Moderate (Grade 2), Severe (Grade 3), Potentially Life Threatening (Grade 4))^14^.

**Table 2.** Local and systemic adverse events: Incidence, frequency and severity by vaccine group and doses.

|  |  |  |  |  |  |  |  |  |
| --- | --- | --- | --- | --- | --- | --- | --- | --- |
| Dose Administration | ARVAC-CG 25-µg Group A |  |  |  | ARVAC-CG 50-µg Group B |  |  |  |
|  | First or second (N = 60) | First (N = 60) | Second (N = 59) | P<sup>A</sup> | First or second (N = 20) | First (N = 20) | Second (N = 18) | P<sup>A</sup> |
| Participants with at least one local adverse event, incidence (frequency) |  |  |  |  |  |  |  |  |
| Any grade p<sup>B</sup> | 46 (76·7) | 41 (68·3) | 28 (47·5) | 0·026 | 12 (60·0)<br>0·1606 | 12 (60·0)<br>0·5874 | 5 (27·8)<br>0·1785 | 0·0585 |
| Grade 1 p<sup>B</sup> | 45 (75·0) | 41 (68·3) | 28 (47·5) | 0·026 | 12 (60·0)<br>0·2555 | 12 (60·0)<br>0·5874 | 4 (22·2)<br>0·0995 | 0·025 |
| Grade 2 p<sup>B</sup> | 7 (11·7) | 5 (8·3) | 2 (3·4) | 0·439 | 2 (10)<br>>0·9999 | 1 (5·0)<br>>0·9999 | 1 (5·6)<br>0·5556 | >0·999 |
| Local adverse events overall frequency, N (%) <sup>C</sup> |  |  |  |  |  |  |  |  |
| Any | 129 (100) | 71 (100) | 58 (100) |  | 34 (100) | 25 (100) | 9 (100) |  |
| Grade 1 | 117 (90·7) | 62 (87·3) | 55 (94·8) |  | 31 (91·2) | 24 (96) | 7 (77·8) |  |
| Grade 2 | 12 (9·3) | 9 (12·7) | 3 (5·2) |  | 3 (8·8) | 1 (4) | 2 (22·2) |  |
| Grade 3 | 0 (0) | 0 (0) | 0 (0) |  | 0 (0) | 0 (0) | 0 (0) |  |
| Grade 4 | 0 (0) | 0 (0) | 0 (0) |  | 0 (0) | 0 (0) | 0 (0) |  |
| Participants with at least one systemic adverse event, N (%) |  |  |  |  |  |  |  |  |
| Any Grade p<sup>B</sup> | 28 (46·7) | 20 (33·3) | 21 (35·6) | 0·848 | 8 (40·0)<br>0·7958 | 8 (40·0)<br>>0·9999 | 4 (22·2)<br>0·3926 | 0·485 |
| Grade 1 p<sup>B</sup> | 28 (46·7) | 19 (31·7) | 21 (35·6) | 0·700 | 8 (40)<br>0·6087 | 7 (35)<br>0·7887 | 4 (22·2)<br>0·3926 | 0·485 |
| Grade 2 p<sup>B</sup> | 6 (10) | 4 (6·7) | 4 (6·8) | >0·99 | 1 (5)<br>0·3282 | 1 (5)<br>>0·9999 | 0 (0)<br>>0·9999 | >0·99 |
| Systemic adverse events overall frequency, N (%) <sup>C</sup> |  |  |  |  |  |  |  |  |
| Any | 101 (100) | 53 (100) | 48 (100) |  | 37 (100) | 25 (100) | 12 (100) |  |
| Grade 1 | 90 (89·1) | 48 (90·6) | 42 (87·5) |  | 34 (91·9) | 22 (88) | 12 (100) |  |
| Grade 2 | 11 (10·9) | 5 (9·4) | 6 (12·5) |  | 3 (8·1) | 3 (12) | 0 (0) |  |
| Grade 3 | 0 (0) | 0 (0) | 0 (0) |  | 0 (0) | 0 (0) | 0 (0) |  |
| Grade 4 | 0 (0) | 0 (0) | 0 (0) |  | 0 (0) | 0 (0) | 0 (0) |  |

Data are expressed as number of individuals, N (percentage, %) or median (interquartile range, IQR).
<sup>A</sup> Exact P Value. First vs. Second dose. Fisher's exact test.
<sup>B</sup> Exact P value. 25-µg vs 50-µg. Fisher's exact test.
<sup>C</sup> Overall adverse events in the same study group.

The most frequent local AE were discomfort/tenderness and pain at injection site in both groups (Figure 2A). All reactions were transient and did not present complications.

Systemic AE were less common. Reported frequency of solicited/unsolicited systemic AEs, was not significantly different between the first and second doses for both groups. In Group A, 33·3% and 35·6% of the volunteers reported at least one systemic AE after the first- and the second administration, respectively (p=0·848), while in Group B, 40·0% and 22·2% of the volunteers reported at least one systemic AE after each dose, respectively (p=0·485) (Table 2).

The most frequent systemic AEs were drowsiness, headache, myalgia, and fatigue (Figure 2B). Only 1 case of fever (38·3°C) lasting 1 day was reported. Of note, 89·9% of reported AE were grade 1 and there was no grade 3 or more severe AE (Table 2).

No abnormal laboratory values were reported to be clinically significant. There were no serious AEs, deaths, or withdrawals due to an AE during the study. No cases of Guillain-Barré syndrome, thromboembolic events, myocarditis or pericarditis, or other AE of special interest have been reported.

### Immunogenicity results

Immune responses presented in this interim report were evaluated after a single ARVAC-CG booster administration. A significant increase of the nAb titres against all the five SARS-CoV-2 VOCs analysed was observed in both ARVAC-CG vaccine cohorts after 14 days, compared to pre-vaccination titres (P<0·0001) (Figure 3). ARVAC-CG 25-μg dose induced a 12·6 (95% CI, 8·8–17·9), 16·6 (95% CI, 11·8–23·4), 11·3 (95% CI, 7·8–16·5), 12·8 (95% CI, 9·2–18·0), and 8·6 (95% CI, 6·1– 12·0) geometric mean fold rise (GMFR) in nAb titres against Ancestral, Gamma, Delta, Omicron BA.1 or Omicron BA.5 variants of SARS-CoV-2, respectively (Figure 3A). Additionally, immunisation with the 50-μg dose induced a GMFR of 29·9 (95% CI, 12·6–70·6), 30·9 (95% CI, 13·4–71·5), 18·4 (95% CI, 8·2–41·1), 29·9 (95% CI, 13·0–68·3), and 13·0 (95% CI, 6·0–28·4) in nAb titres against Ancestral, Gamma, Delta, Omicron BA.1 or Omicron BA.5 variants of SARS-CoV-2, respectively (Figure 3B). Of note, GMFR of nAb titres against Ancestral and Omicron BA.1 variants were significantly higher in Group B than in Group A (P=0·0448 and P=0·0271, respectively) (Appendix table S2).

**Figure 3:**
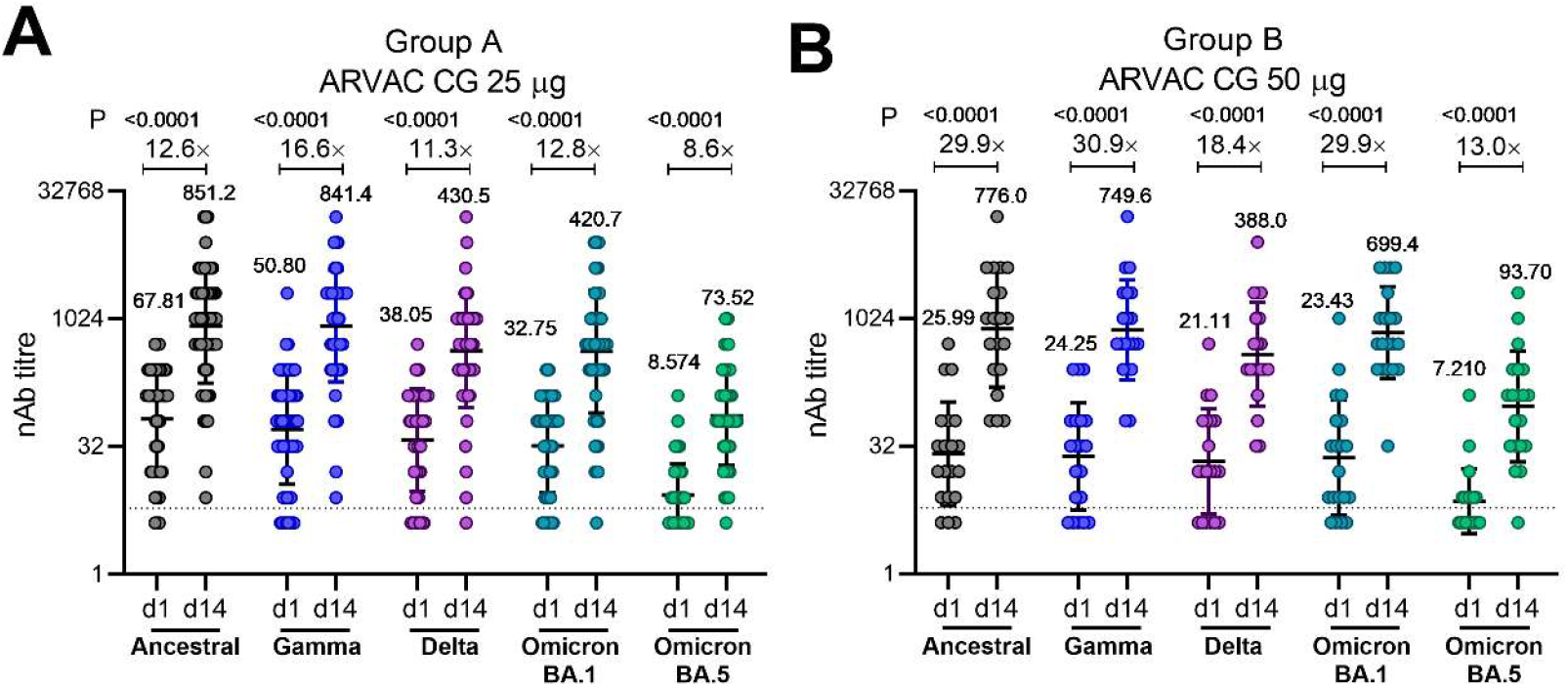
Administration of ARVAC-CG booster increases the nAb titres against the Ancestral, Gamma, Delta, Omicron BA.1 and Omicron BA.5 variants of SARS-CoV-2. The nAb titres against the Ancestral, Gamma, Delta and Omicron BA.1 and Omicron BA.5 variants of SARS-CoV-2 in plasma samples of individuals boosted with ARVAC-CG 25μg (A) or 50 μg (B) prior to the vaccine administration (d1) or after 14 days of booster administration (d14). Each point represents the nAb titre of a volunteer at the indicated time point and against the depicted viral variant. The nAb geometric mean titres (GMTs) and 95% CIs are shown as horizontal and error bars, respectively. The numbers depicted above the individual points for each specified time point and viral variant represent de GMT. The fold increases in the GMT from day 1 to day 14 (GMFR) for each specified variant are shown with a number followed by a ×. The dashed line represents the positivity threshold on the Neutralisation assay. Statistical differences were analysed using the Wilcoxon pair-matched test. Exact P values are depicted above the data sets that were compared.

After 14 days of a booster with ARVAC-CG the 4×-seroconversion rates for the Ancestral SARS-CoV-2 strain were 88·3 % (95% CI, 77·8–94·2) in the 25-μg cohort and 90·0% (95% CI, 69·9–98·2) in the 50-μg cohort. Whereas the corresponding 4×-seroconversion rates were 90·0% (95% CI, 79·8– 95·3) and 85% (95% CI, 64·0–94·8) against the Gamma VOC, 80·0% (95% CI, 68·2–88·2) and 85·0% (95% CI, 64.0–94.8) for Delta VOC, 93·3% (95% CI, 84·1–97·4) and 85·0% (95% CI, 64·0–94·8) for Omicron BA.1 VOC, and 80·0% (95% CI, 68·2–88·2) and 80.0% (95% CI, 58·4–91·9) for Omicron BA.5 VOC, respectively (Appendix table S2).

Fourteen days after boosting, 4×-seroconversion rates for all tested variants were similar in both groups, while 10×-seroconversion rates for Omicron BA.1 VOC were significantly higher in the 50-μg cohort (Appendix table S2).

Based on pre-existing anti-N IgG titres and/or previous history of COVID-19, individuals were stratified into two populations: seronegative individuals with no previous COVID-19 history and seropositive and/or with previous history of COVID-19 individuals. Both populations developed similar nAb GMTs against Ancestral, Gamma, Delta, Omicron BA.1 and Omicron BA.5 after 14 days of ARVAC GC booster. Moreover, GMFR from baseline were similar for groups A and B (Appendix Figure S1).

No significant differences were found in nAb responses and seroconversion rates between female or male volunteers, except for the 4×-seroconversion rate in nAbs against Delta VOC (Appendix table S3).

When volunteers were subdivided regarding the primary vaccination received, a significant increase in the nAb GMTs against the different viral variants was observed in all previously vaccinated subgroups (Figure 4).

**Figure 4:**
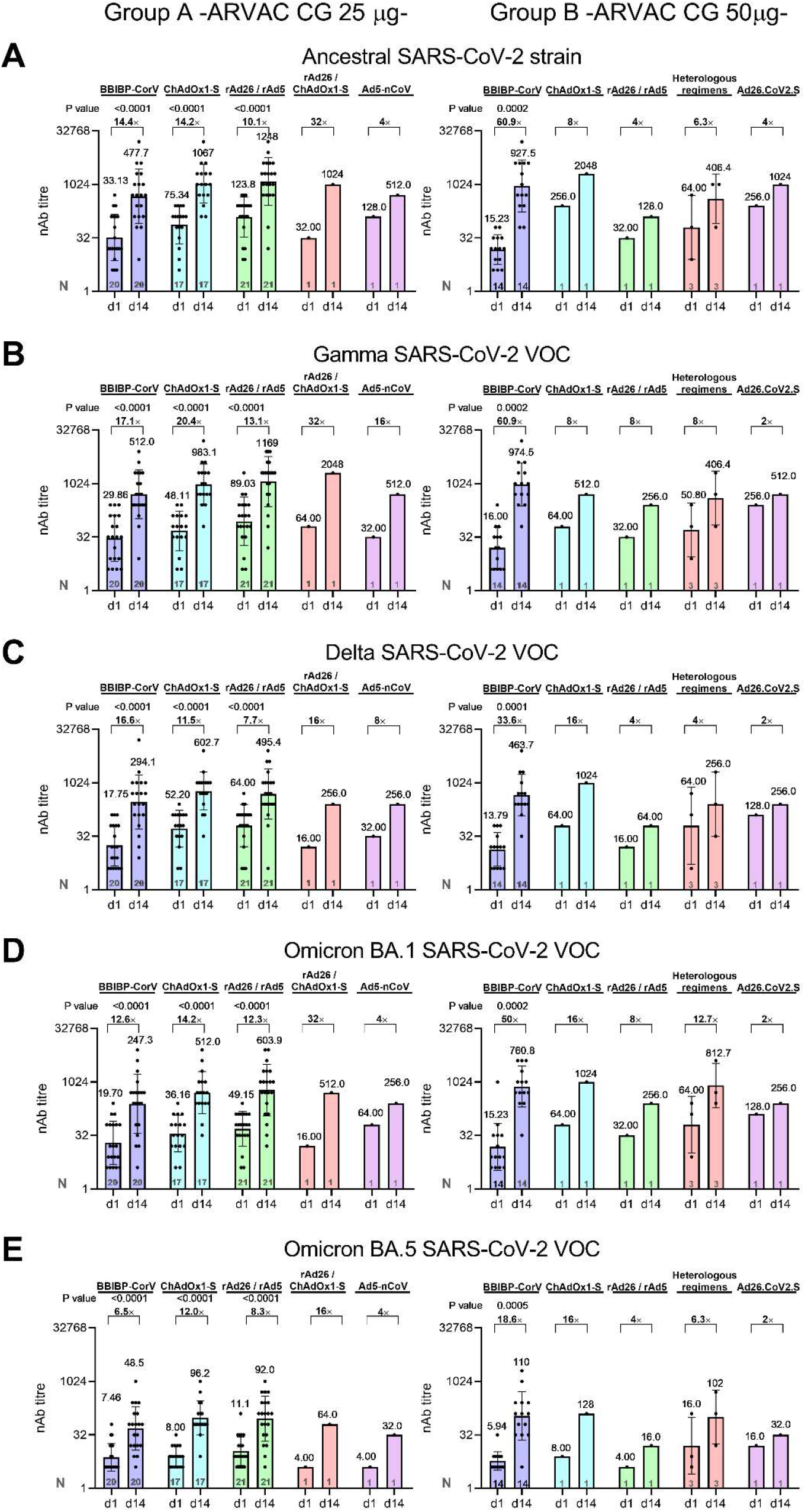
Administration of ARVAC-CG as booster increases the nAb titres against the Ancestral, Gamma, Delta, Omicron BA.1 and Omicron BA.5 variants of SARS-CoV-2 in individuals with different primary vaccinations schemes. Neutralising antibody titres against the Ancestral (A), Gamma (B), Delta (C), Omicron BA.1 (D) and Omicron BA.5 (E) variants of SARS-CoV-2 in plasma samples of individuals boosted with ARVAC-CG 25μg (left panels)) or 50 μg (right panels) prior to the vaccine administration (d1) or after 14 days of booster administration (d14). Participants in each cohort were grouped according to the primary vaccination scheme received (BBIBP-CorV (Sinopharm), ChAdOx1-S (Oxford AstraZeneca), rAd26/rAd5 (Sputnik V, Gamaleya Institute), Ad5-nCoV (CanSino), Ad26.CoV2.S (Janssen), rAd26/ChAdOx1-S (heterologous vaccination regime Sputnik V Component 1/ Oxford AstraZeneca) or heterologous vaccination regimens (ChAdOx1-S / mRNA1273 or BBIBP-CorV / BNT162b2). Each point represents the nAb titre of a volunteer at the indicated time point and against the depicted viral variant. The nAb GMTs with 95% CIs are shown as horizontal and error bars, respectively. The numbers depicted above the individual points for each specified time point and viral variant represent GMT values. The fold increases in GMT values from day 1 to day 14 (GMFR) for each specified variant are shown with a number followed by a ×. The dashed line represents the positivity threshold on the Neutralisation assay. Statistical differences were performed using the Wilcoxon pair-matched test. Exact P values are depicted above the data sets that were compared.

Similar results were observed at 28 days post-booster vaccination with ARVAC-CG (Appendix Figure S2 and table S4). Levels of anti-RBD and anti-spike antibodies raised significantly after 28 days with respect to baseline levels in both study groups (Appendix Figure S3).

To preliminarily assess the comparative immunogenicity of ARAVAC GC, nAb GMTs and GMFR were compared with those in samples from a cohort of individuals out of the protocol, with similar demographic characteristic, who had received a heterologous booster dose with the Ancestral-based BNT16b2 mRNA vaccine. Baseline nAb GMTs against the five viral variants in both ARVAC-CG cohorts were similar to those observed before the booster with BNT16b2 (P>0·05) (Figure 5A-E). After 14 or 28 days the nAb GMTs against the Ancestral SARS-CoV-2 in group A were similar to that achieved after BNT16b2 booster (Figure 5A). However, nAb GMTs against Gamma, Delta, Omicron BA.1, and Omicron BA.5 were significantly higher in group A than those reached in BNT16b2 boosted individuals (Figure 5B-E). Similarly, in group B nAb GMTs against Gamma VOC after 14 days of booster and against Omicron BA.1 and Omicron BA.5 VOCs at all tested timepoints were significantly higher to the corresponding nAb GMTs in BNT16b2 boosted individuals (Figure 5B, D, and E). While GMFR in nAb titres against Ancestral, Gamma and Delta VOCs after a booster dose with BNT16b2 were similar to those elicited in the ARVAC-CG cohorts (P>0.05), GMFR of nAb titres against Omicron BA.1 and BA.5 were significantly lower in the BNT16b2-than in the ARVAC-CG boosted individuals (Figure 5F).

**Figure 5:**
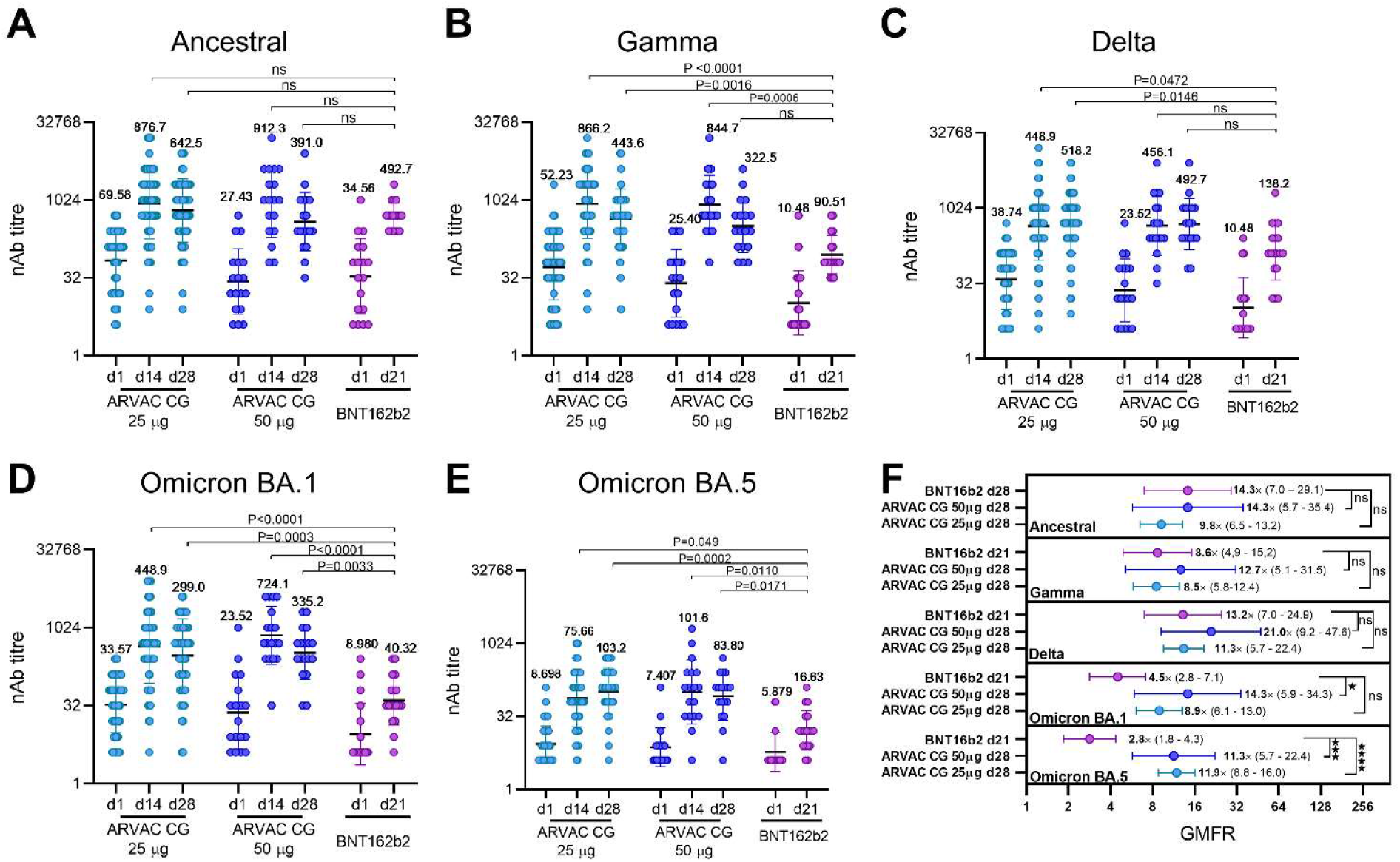
Comparison of nAb GMT and GMFR after booster with ARVAC-CG or booster with BNT16b2 in a contemporaneous boosted study. Neutralising antibody titres against the Ancestral **(A)**, Gamma **(B)**, Delta **(C)**, Omicron BA.1 **(D)** and Omicron BA.5 **(E)** variants of SARS-CoV-2 in plasma samples of individuals boosted with the indicated vaccine (ARVAC-CG 25μg, ARVAC-CG 50 μg or BNT16b2) prior to the vaccine administration (d1) or at the indicated days after booster administration (d14, d21, d28). Each point represents the nAb titre of a volunteer boosted with the indicated vaccine, at the indicated time point and against the depicted viral variant. The nAb GMTs and 95% CIs are shown as horizontal and error bars, respectively. The numbers depicted above the individual points for each specified time point and viral variant represent the GMTs. Statistical differences were analysed using the Kruskal-Wallis test followed by the Dunn’s multiple comparison test. Exact P values are depicted above the data sets that were compared. ns: P>0.05 **(F)** Fold increases in the GMT from day 1 to day 21 or 28 (GMFR) for each specified variant represented by a point and written with a number followed by a ×. The horizontal lines represent the 95% CIs. Statistical differences were analysed using Kruskal-Wallis test followed by the Dunn’s multiple comparison test. ns: P>0·05, ⋆ : P<0·05; ⋆ ⋆ ⋆ : P<0·001; ⋆ ⋆ ⋆ ⋆ : P<0·0001

Transformation of nAb titres to IU/ml, allowed the evaluation of the percentage of participants with nAb levels associated with high VE. In group A, rates of participants with nAb levels higher than of 947 IU/ml before the booster were 26·7 % (95% CI 39·0, 17·1), 3·3 % (95% CI 11·4, 0·6), 6·7 % (95% CI 15·9, 2·6), and 1·7 % (95% CI 8·9, 0·1) for the Ancestral, Gamma, Delta, and Omicron BA.1 SARS-CoV-2 VOCs, respectively. These rates raised significantly after 14 days of having received the booster to reach 83·3 % (95% CI 90·7, 72·0), 80·0 % (95% CI 88·2, 68·2), 60·0 % (95% CI 71·4, 47·4), and 78·3 % (95% CI 86·9, 66·4) for each viral variant, respectively. Likewise, in group B the proportion of participants with nAb levels higher than 947 IU/ml raised significantly from 15·0 % (95% CI 5·2, 36·0), 0·0 % (95% CI 0·0, 16·1), 5·0 % (95% CI 0·3, 23·6), and 10·0 % (95% CI 1·8, 30·1) for the Ancestral, Gamma, Delta, and Omicron BA.1 SARS-CoV-2 VOCs respectively at baseline, to 80·0 % (95% CI 58·4, 91·9), 70·0 % (95% CI 48·1, 85·5), 45·0 % (95% CI 25·8, 65·8), and 95·0 % (95% CI, 76·4, 99·7) for each viral variant, respectively. (Appendix Figure S4A-B).

A significant increase in the frequency of IFN-γ producing cells upon *in vitro* re-stimulation with SARS-CoV-2 RBD peptide pools was observed in both ARVAC-CG cohorts in comparison with the levels observed before the booster dose. In addition, a slight increase of IL-4 producing cells was observed after the booster in group A participants (Figure 6). Interestingly, booster vaccination with - 25-μg or 50-μg of ARVAC-CG led to increases in antigen-specific cellular immune responses in individuals primed with different vaccine platforms (Appendix Figure S5).

**Figure 6.**
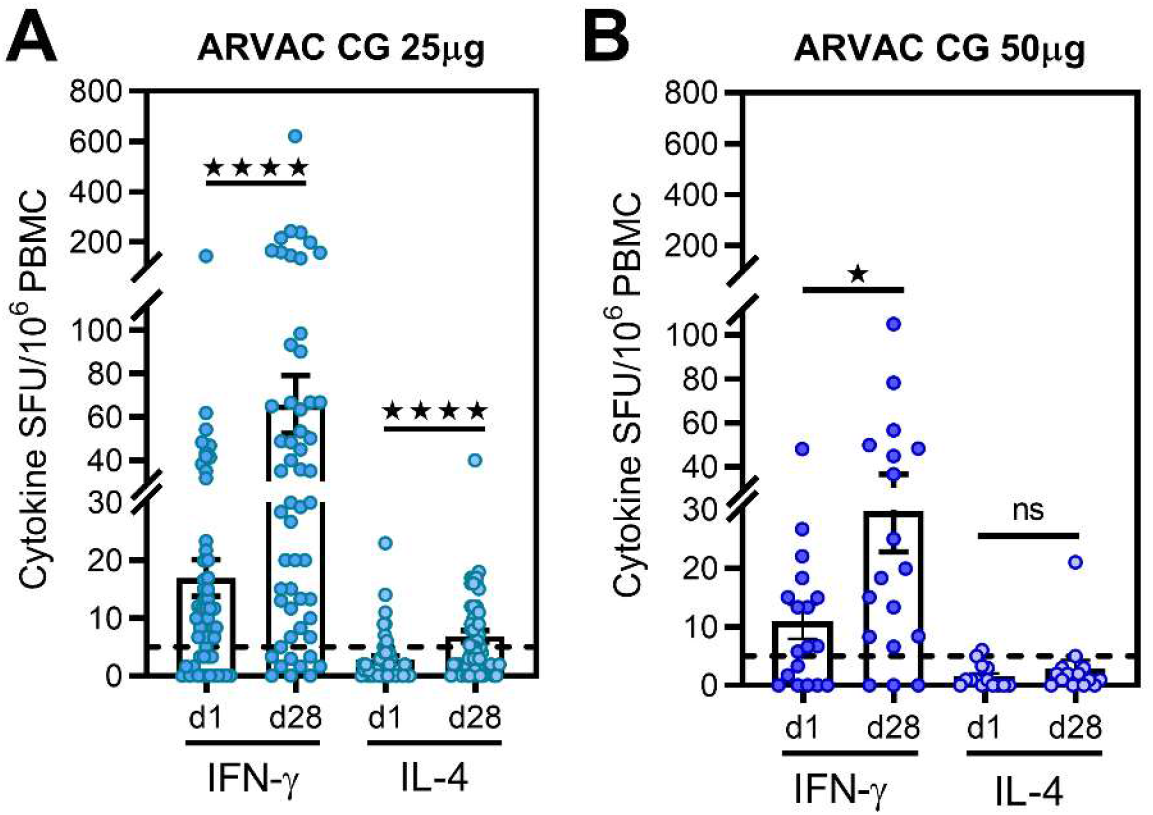
ARVAC-CG booster induces significant increase of Th1-predominant cell response measured by IFN-γ and IL-4 ELISpot after restimulation of PBMCs with RBD spanning peptide pool. Before booster administration (1d) and after 28 days (d28) of administration of ARVAC-CG 25 μg (A) or 50 μg (B) dose, RBD specific cellular responses were measured by IFN-γ and IL-4 ELISpot in PBMCs. Shown are spot-forming units (SFU) per 1 × 10^6^ PBMCs producing IFN-γ and IL-4 after stimulation with-RBD peptide pool. Bars indicate the mean, and error bars the SEM. Statistical analysis was performed by the Wilcoxon matched pairs signed rank test. ⋆ ⋆ ⋆ ⋆ : P < 0·0001; ⋆ : P<0·05; ns: P>0·05.

## Discussion

The main findings of this study are that the vaccine candidate ARVAC-CG when given as a booster dose is well tolerated and induces a robust and broad nAb response against several SARS-CoV-2 VOCs.

The interim results of this phase 1 study indicate that ARVAC-CG vaccine given to individuals who previously received a complete primary vaccination regimen has a clinically acceptable safety and reactogenicity profile for both antigen doses (25-μg and 50-μg).

Immunogenicity results indicate that ARVAC-CG as a booster dose induces a sharp increase of broadly nAb titres against the Ancestral SARS-CoV-2 strain, the Gamma SARS-CoV-2 VOC, which is the vaccine prototype strain, as well as against antigenic distant SARS-CoV-2 VOCs like Delta, Omicron BA.1 and Omicron BA.5. Moreover, the ARVAC-CG 50-μg dose outperformed the 25-μg dose in terms of nAb titres reached against the Ancestral and Omicron BA.1 viral variants and 10×-seroconversion rates in nAb against Omicron BA.1. The booster effect of ARVAC-CG vaccine was evident despite the variety of immunisation schemes received by the per protocol included population.

With the emergence of new VOCs it is clear that breakthrough infections can occur in vaccinated persons, including those with previous SARS-CoV-2 infection.^17,18^ Therefore, the capability to boost the immune responses in individuals with a previous history of infection becomes critical, enhancing protection against COVID-19 and post-COVID conditions.^19^ The results of this study highlight the potential benefit of the ARVAC-CG vaccine for all populations regardless of their prior COVID-19 serological status.

The performance of the ARVAC-CG booster dose regarding nAb GMTs and GMFR, was similar to that of the BNT16b2 booster against Ancestral and Delta VOCs, but better for Gamma, Omicron BA.1 and BA.5 VOCs. Similarly, booster shots with Beta variant-based vaccines elicit broad nAb responses against the Ancestral, the Beta and the Omicron BA.1 VOC, which were higher in terms of nAb GMT or GMFR than that elicited by the Ancestral prototype vaccines, or a homologous booster with BNT16b2.^10,20,21^ Results presented in this work suggest that a booster with ARVAC-CG increases the breath of the nAb and are in agreement with non-clinical results of this vaccine formulation^13^.

Even though there are not well-established threshold values of nAb levels that correlate with protection against symptomatic and asymptomatic SARS-CoV-2 infection, an increasing number of studies that use standardised methods offer hints about the nAb levels that might be associated with protection.^18,22-24^ In this regard, nAb levels of ∼100-120 IU/ml have been correlated with ∼80% VE against symptomatic infection, whereas nAb levels of ∼900-1030 IU/ml correlate with ∼90% VE against symptomatic infection.^22^ Results presented here suggest that a booster with ARVAC-CG increases significantly the proportion of individuals with nAb titres that correlate with high VE.

T-cell immunity is crucial to combat acute SARS-CoV-2 infection and for the development of long-term immunity.^25^ While antibody titres tend to wane rapidly and show limited neutralising activity to newly arising VOCs, T-cell memory is largely conserved.^25^ ARVAC-CG boosted T cell immunity, which might contribute to eliminate virus-infected cells. Although it has been reported that durable spike-specific T-cell responses after different COVID-19 primary vaccination regimens are not further enhanced by an mRNA booster,^26^ results presented here show that ARVAC-CG booster significantly increases the proportion of antigen-specific IFN-γ and IL-4 producing T cells in individuals previously vaccinated with different primary schemes.

One limitation to this study is the lack of randomization for volunteers’ recruitment. Nevertheless, despite the sequential study design, enrolment dates of the booster groups occurred within weeks of each other, representing similar epidemiologic environments of circulating variants.

In this phase 1 study, safety was demonstrated after two ARVAC-CG administrations highlighting that ARVAC-CG is safe. Immune responses presented were assessed only after 14 and 28 days of one booster administration, however longer-term follow-up of immune responses will have to be studied in a phase 2/3 study.

While both formulations of ARVAC-CG exhibited a favourable safety and reactogenicity profile eliciting broadly nAb responses and T cell immunity against SARS-CoV-2, the 50-μg dose outperformed the 25 -μg dose in certain of the immunogenicity variables evaluated. Therefore, it might be appropriate to test the 50-μg dose vaccine in a Phase 2/3 study to evaluate its safety, and immunogenicity in a larger population. Selection of the 50-μg vaccine dose may allow the testing of a bivalent vaccine containing 25-μg of Gamma-based antigen plus 25-μg of Omicron BA.5-based antigen to improve the response capacity, as part of a Phase 2/3 study, in accordance with the recommendations from the advisory committee on immunisation practices for the use of bivalent booster doses of COVID-19 vaccines to increase protection against circulating VOCs and to broaden neutralisation to previous and potentially yet-to-emerge variants.^27^

There is a need for widespread immunisation programs with new generation of COVID-19 booster vaccines which provide a wide breadth of protection against constantly emerging SARS-Cov-2 variants. Nonetheless, LMICs lag far behind in this effort due to limitations in affordability and accessibility to vaccines. Hence, developments such as ARVAC GC may offer an opportunity to overcome some of these challenges and improve response capacity of many countries, worldwide.

## Data Availability

All data produced in the present work are contained in the manuscript.
Anonymised individual participant data will be made available when the study is complete, on reasonable requests made to the corresponding author. Proposals will be reviewed and approved by the sponsor, investigator, and collaborators on the basis of scientific merit. After approval of a proposal, data can be shared through a secure online platform. All data will be made available for a minimum of 5 years from the end of the trial.

## Laboratorio Pablo Cassará R&D and CMC group

Sabrina A del Priore; Andrés C. Hernando Insua, Ingrid G. Kaufmann; Adrián Di María; Adrián Góngora; Agustín Moreno; Susana Cervellini; Blasco Martin; Esteban Ali; Romina Albarracín; Barsanti Bruno; Fernando Toneguzzo, Guillermina Sasso, Sebastian Stamer; Regina Cardoso and Alejandro Chajet.

## Contributors

JC, ML, JCV, FMO, JMR, JF, KAP, and LMC conceptualized and designed the study. KAP, LMC and JC wrote and edited the manuscript. ML, FMO and JMR were responsible for project administration. JC, KAP, LMC, ML, and JF, accessed, verified, and analysed the data, and generated the interim report. EF is the study site scientific director and supervised the study. GAY is a study site principal investigator. KH, GAY, and PEPL were responsible for the site work including the recruitment, follow up, and data collection. BM, MS, AD, CPC, LS, LB performed immunogenicity experiments. AC, AV, and JG contributed to the performance and analysis of neutralizing antibody assays. SG, VVGM, and EE provided the plasma samples from individuals boosted with authorized vaccines. JMR, JCV, FMO and LPC R&D and CMC Group were responsible for R&D for chemistry, manufacture and controls of antigen and study products. JCV and FMO provided regulatory oversight. ML was responsible for the overall supervision of the study and monitored the trial. All authors contributed to data interpretation, review, and editing of this manuscript. All authors have read and approved the final version of the manuscript.

## Acknowledgments

We thank the trial volunteers for their contribution and commitment to vaccine research. We are thankful to the staff of Laboratorio Pablo Cassará for their essential contribution to the development, scaling, manufacture, control and stability study of the clinical batches of ARVAC-CG antigen and vaccine: Krum, Valeria; Drehe, Ignacio; Zurvarra, Francisco; Baque, Jonathan; Payes, Cristian; Heinrich, Brenda; Gambone, Melisa; Descoins, Horacio; De Nichilo, Analía; Sidabra, Johanna; Licausi, Mariana; Cortez, Christian; Roman, María Victoria; Villar, Alejandra; Diaz, Sebastián; Frattolillo, Matías; Mestre, Diego; Gonzalo, Javier; Maluvini, Ernesto; Sperandini, Cecilia; Farre, Paola; Trovato, Fernando; Strada, Ariel; Cocciolo, Giovanna; and Privitera, Anabella. We additionally thank Roberto Debagg for giving valuable advice during to the design and completion of the clinical trial. We thank Daniela Hozbor for the access to the samples obtained during the COVID-19 serology surveillance strategy implemented by the Ministry of Health of the Province of Buenos Aires. We thank Florencia Quiroga, Gabriela Turk and Natalia Laufer for performing T cell stimulation assays by ELISpot at Instituto de Investigaciones Biomédicas en Retrovirus y SIDA, INBIRS-CONICET, Buenos Aires, Argentina. We also thank Jaime Torres who provided medical writing support. We thank Elisa Estenssoro of the Ministry of Health of the Province of Buenos Aires for critical review of the manuscript. We are grateful to Dr. Ángela Spanguolo de Gentile, Dr. Pablo Bonvehí, and Dr. Hugo Krupitzki who are members of the external Independent Committee of Data Review for their dedication and continuous invaluable advice.

