## Supplementary Material for "Safety, tolerability, and immunogenicity of a new SARS-CoV-2 recombinant Gamma variant RBD-based protein adjuvanted vaccine, used as heterologous booster in healthy adults: a Phase 1 interim report"

### Supplementary Figures and Tables

**Figure S1**

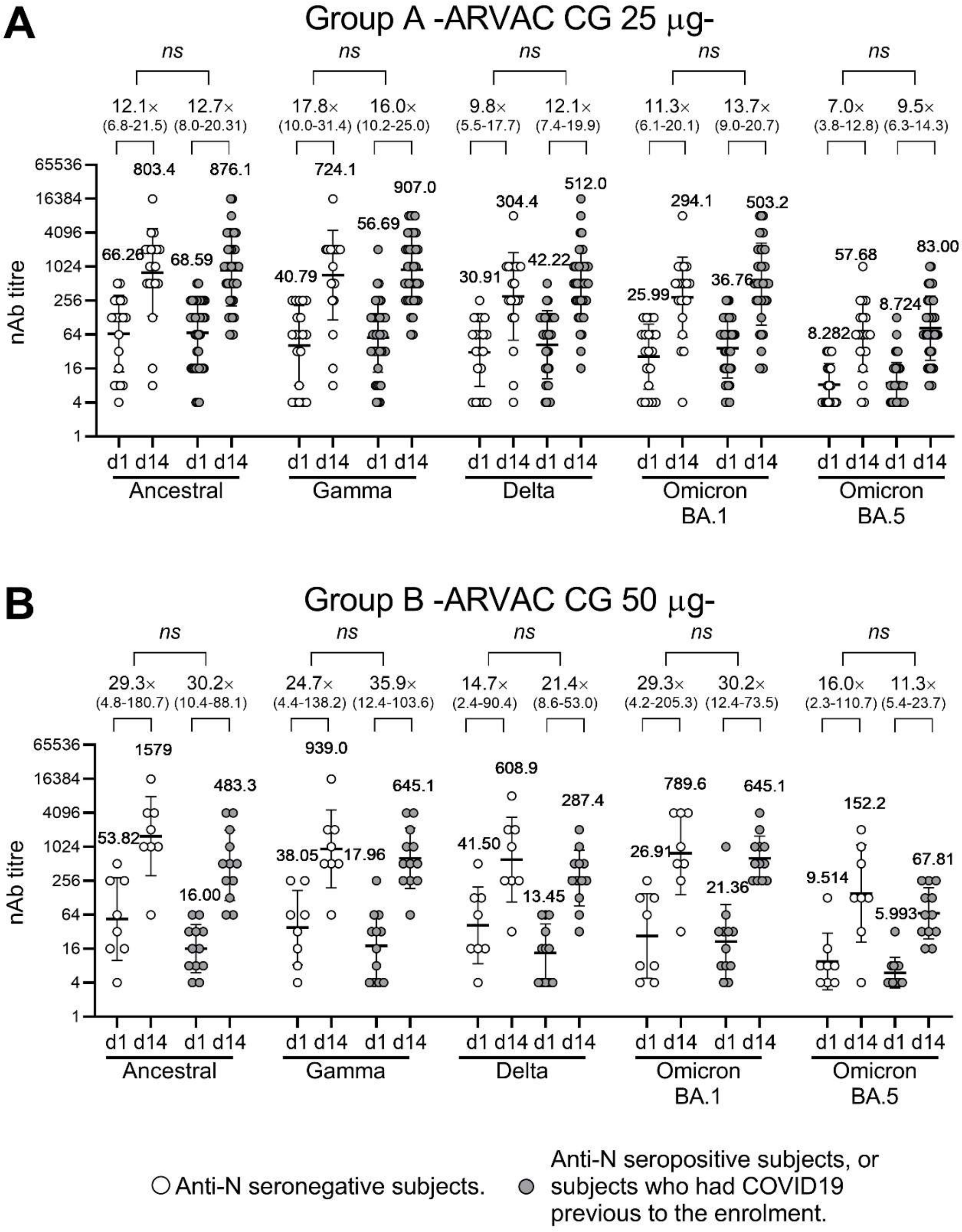


**Figure S1: Administration of ARVAC-CG as booster increases the Neutralisation antibody titres against the Ancestral, Gamma, Delta, Omicron BA.1 and Omicron BA.5 variants of SARS-CoV-2 irrespective of the previous history of COVID19 or anti-N serology of the study participants.** Study participants were classified according to their previous story of COVID19 and/or their anti-N serology into two groups: in the graphs are shown with open circles those who were seronegative for anti-N IgG (at day 1 and at day 28) and have no previous diagnostic of COVID19 and shown with filed circles are those who were seropositive for anti-N IgG (at day 1 or at day 28) or have had COVID19 previous to the study. The neutralising antibody titres against the Ancestral, Gamma, Delta and Omicron BA.1 and Omicron BA.5 variants of SARS-CoV-2 in plasma samples of individuals boosted with ARVAC-CG 25µg (A) or 50 µg (B) prior to the vaccine administration (d1) or after 14 days of booster administration (d14) are shown. Each point represents the nAb titre of a volunteer at the indicated time point and against the depicted viral variant. The nAb geometric mean titres (GMTs) with 95% CIs are shown as horizontal and error bars, respectively. The numbers depicted above the individual points for each specified subgroup, time point and viral variant represent the GMT. The fold increases in the GMT from day 1 to day 14 (GMFR) for each specified subgroup of participants and variant are shown with a number followed by a × with the 95% CI written bellow between brackets. Statistical differences were analysed using the Mann Whitney test. ns: P>0.05.

**Supplementary Figure 2**

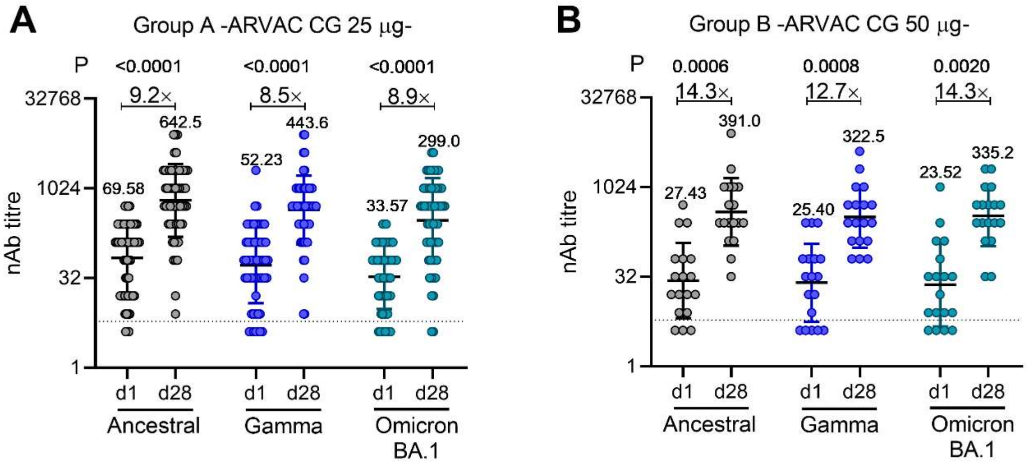


**Figure S2: Administration of ARVAC-CG as booster increases after 28 days the nAb titres against the Ancestral, Gamma and Omicron BA.1 variants of SARS-CoV-2.** Neutralising antibody titres against the Ancestral, Gamma, and Omicron BA.1 variants of SARS-CoV-2 in plasma samples of individuals boosted with ARVAC-CG 25µg (A) or 50 µg (B) prior to the vaccine administration (d1) or after 28 days of booster administration (d28). Each point represents the nAb titre of a volunteer at the indicated time point and against the depicted viral variant. The nAb geometric mean titres (GMT) with 95% CIs are shown as horizontal and error bars, respectively. The numbers depicted above the individual points for each specified time point and viral variant represent the GMT. The fold increases in the GMT from day 1 to day 28 (GMFR) for each specified variant are shown with a number followed by a ×. The dashed line represents the positivity threshold on the Neutralisation assay. Statistical differences were analysed using the Wilcoxon pair-matched test. Exact P values are depicted above the data sets that were compared.

Supplementary Figure 3

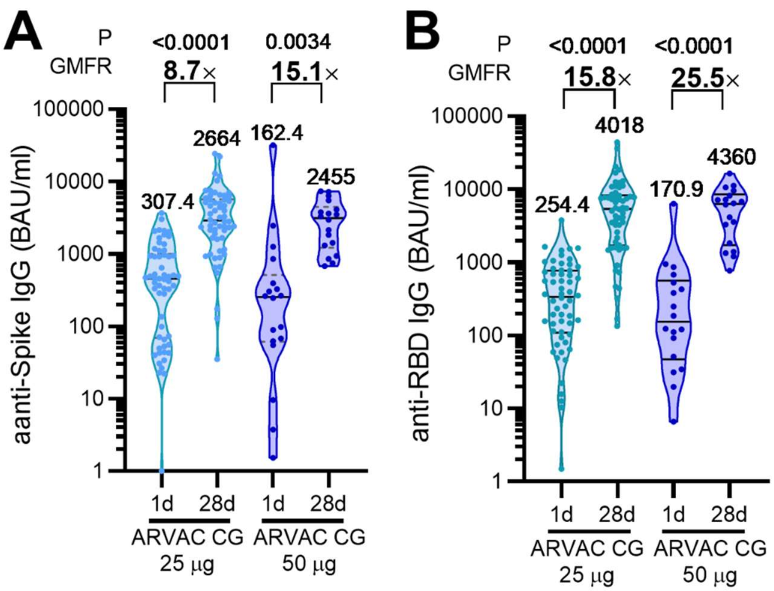


**Figure S3. Analysis of binding antibody response induced by ARVAC-CG booster dose.** (A) Serum anti-spike IgG (A) or anti-RBD IgG (B) were analysed by ELISA. Antibody levels are expressed in BAU/ml according to the WHO International Antibody Standard. Graphs display violin plots showing the frequency distribution of the data and dots show individual values for each volunteer at a specified time point. The geometric means are shown above the violin plots. The fold raise in GM (GMFR) after 28 days of booster (d28) respect to baseline (d1) is shown above a line connecting both time points. The Wilcoxon matched pairs test was used for statistical analysis. P values are depicted on the graph. Ns: P>0.05.

**Supplementary Figure 4**

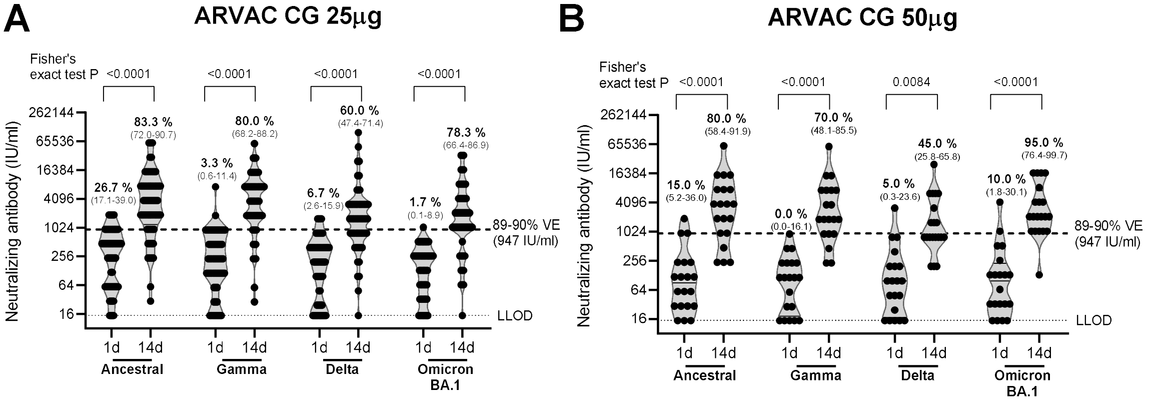


**Figure S4: Administration of a booster dose of ARVAC-CG increases the frequency of individuals with nAb levels that correlate with high VE.** The nAb titres prior and after the booster were transformed to international units per ml (IU/ml) by the inclusion in each plate of a secondary standard that was calibrated with the WHO international standard (NIBSC code: 20/268). A cut off value 947 IU/ml was used to determine the proportion of individuals with nAb levels ≥ 947 IU/ml (levels associated with 80-90% VE) prior to the booster administration (d1) or 14 days after administration (d14). Each point represents the nAb level (IU/ml) of a volunteer at the indicated time point and against the depicted viral variant. The geometric mean of nAb levels with 95% CIs are shown as horizontal and error bars, respectively. The numbers depicted above the individual points for each specified time point and viral variant represent the percentage of individuals with nAb levels ≥ 947 IU/ml and the respective 95% CI. The dashed line represents the positivity threshold on the Neutralisation assay (LLOD). Statistical differences were analysed using the Fisher´s exact test. Exact P Values are depicted above the data sets that were compared.

**Supplementary Figure 5**

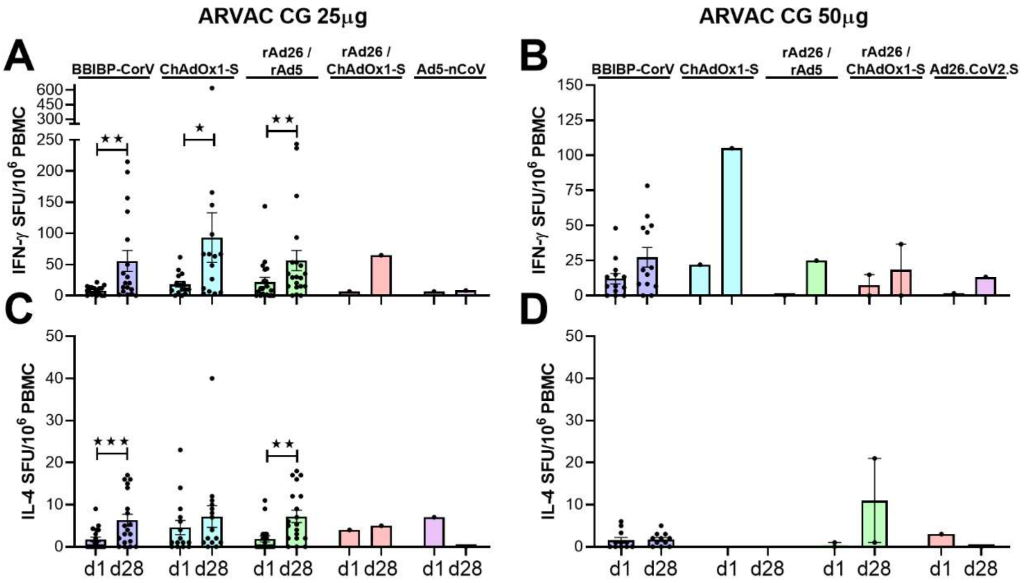


**Figure S5. ARVAC-CG booster induces significant increase of Th1-predominant cell response measured by IFN-γ and IL-4 ELISpot after restimulation of PBMCs with RBD spanning peptide pool in individuals previously vaccinated with different primary vaccination schemes.** Before booster administration (1d) and after 28 days (d28) of administration of ARVAC-CG 25 µg (A, C) or 50 µg (B, D) dose, RBD-specific cellular responses were measured by IFN-γ (A, B) and IL-4 (C, D) ELISpot in PBMCs. Shown are spot-forming units (SFU) per $1 \times 10^6$ PBMCs producing IFN-γ and IL-4 after stimulation with-RBD peptide pool. Participants in each cohort were grouped according the received primary vaccination scheme (BBIBP-CorV (Sinopharm), ChAdOx1-S (Oxford AstraZeneca), rAd26/rAd5 (Sputnik V, Gamaleya Institute), Ad5-nCoV (CanSino), Ad26.CoV2.S (Janssen), rAd26/ChAdOx1-S (heterologous vaccination regime Sputnik V Component 1/ Oxford AstraZeneca) or heterologous vaccination regimens (ChAdOx1-S / mRNA1273 or BBIBP-CorV / BNT162b2). Each point represents the cytokine spot forming units (SFU) at the indicated time point. The SFU mean is represented by bars and SEM by error bars, respectively. Statistical differences were performed using the Wilcoxon pair-matched test. Ns: P>0.05; ★: P<0.05; ★★: P<0.01, ★★★: P<0.001.

**Supplementary tables**

**Table S1**

| **Table S1: Demographic Characteristics of the Participants in the observational booster study with BNT16b2.** | | |
|---|---|---|
| Variable | | **ARVAC-CG 25-µg Group A** |
| No. of participants | | 18 |
| Sex | Men (N (%)) | 5 (27·8) |
| | Women (N (%)) | 13 (72·2) |
| Age (Median (IQR)) | | 36·5 (32-45) |
| COVID-19 Primary Vaccine Platform | Sinopharm (N (%)) | 2 (11·1) |
| | Sputnik V (N (%)) | 16 (88·9) |
| | Time since last dose of primary immunisation schedule (in months, (Median (IQR)) | 4.2 (3·8-4·6) |
| Prior COVID-19 | No (N (%)) | 18 (100·0) |
| | Yes (N (%)) | 0 (0·0) |

### Table S2

**Table S2: Neutralising antibody and seroconversion analysis against Ancestral SARS-CoV-2 (Wuhan), Gamma, Delta, Omicron BA.1 and Omicron BA.5 after 25 or 50 µg of ARVAC-CG as Booster Dose in Participants.**

| **SARS-CoV-2 variant** | **Ancestral (Wuhan)** | | **Gamma** | | **Delta** | | **Omicron BA.1** | | **Omicron BA.5** | |
|---|---|---|---|---|---|---|---|---|---|---|
| **ARVAC-CG Dose** | **25 µg** | **50 µg** | **25 µg** | **50 µg** | **25 µg** | **50 µg** | **25 µg** | **50 µg** | **25 µg** | **50 µg** |
| No. of participants evaluated[a] | (N=60) | (N=20) | (N=60) | (N=20) | (N=60) | (N=20) | (N=60) | (N=20) | (N=60) | (N=20) |
| **Before booster** | | | | | | | | | | |
| **GMT[b]** | **67·81** | **25·99** | **50·8** | **24·25** | **38·05** | **21.11** | **32.75** | **23.43** | **8·574** | **7.21** |
| (95% CI)[c] | (46·88-98·07) | (13·46-50·2) | (34·6-74·58) | (12·32-47·74) | (26·57-54·49) | (10.81-41.22) | (23.63-45.38) | (11.26-48.75) | (6·895-10·66) | (4.778-10.88) |
| **Day 14** | | | | | | | | | | |
| **GMT[b]** | **851·2** | **776** | **841·4** | **749·6** | **430·5** | **388** | **420.7** | **699.4** | **73·52** | **93.7** |
| (95% CI)[c] | (569·3-1273·0) | (370·4-1626·0) | (568·8-1245·0) | (397·0-1415·0) | (288·4-642·6) | (200.4-751.2) | (273.3-647.5) | (391.2-1250.0) | (52·0-104·0) | (46.4-189.1) |
| **GMFR[d]** | **12·6** | **29·9** | **16·6** | **30·9** | **11·3** | **18.4** | **12.8** | **29.9** | **8·6** | **13.0** |
| (95% CI)[c] | (8·8-17·9) | (12·6-70·6) | (11·8-23·4) | (13·4-71·5) | (7·8-16·5) | (8.2-41.1) | (9.2-18.0) | (13.0-68.3) | (6·1-12·0) | (6.0-28.4) |
| Mann Whitney test P | 0·0448 (*) | | 0·1108 (ns) | | 0·1484 (ns) | | 0·0297 (*) | | 0·2524 (ns) | |
| **4× Seroconversion at day 14[e]** | | | | | | | | | | |
| **Percentage of participants (95% CI)** | **88·3** (77·8-94·2) | **90·0** (64·0-94·8) | **90·0** (79·9-95·3) | **85·0** (64·0-94·8) | **80·0** (68·2-88·2) | **85.0** (64.0-94.8) | **93.3** (84.1-97.4) | **85.0** (64.0-94.8) | **80·0** (68·2-88·2) | **80.0** (58.4-91.9) |
| Fisher's exact test, P | >0,9999 (ns) | | 0·6836 (ns) | | 0·7498 (ns) | | 0·3582 (ns) | | >0·9999 (ns) | |
| Chi-square test, P | 0·8381 (ns) | | 0·5400 (ns) | | 0·6198 (ns) | | 0·2534 (ns) | | >0·9999 (ns) | |
| **10× Seroconversion at day 14[f]** | | | | | | | | | | |
| **Percentage of participants (95% CI)** | **45·0** (33·1-57·5) | **70·0** (48·1-85·5) | **61·7** (49·0-72·9) | **70·0** (48·1-85·5) | **40·0** (28·6-52·6) | **65.0** (43.3-81.9) | **48.3** (36.2-60.7) | **75.0** (53.1-88.8) | **38·3** (27·1-51·0) | **55.0** (34.2-74.2) |
| Fisher's exact test, P | 0·0715 (ns) | | 0·5970 (ns) | | 0·0707 (ns) | | 0·0427 (*) | | 0·2049 (ns) | |
| Chi-square test, P | 0·0527 (ns) | | 0·5020 (ns) | | 0·0521 (ns) | | 0·0379 (*) | | 0·1916 (ns) | |

[a] The number of participants with nonmissing data at baseline or at 28 days in shown.

[b] GMT: Geometric Mean titre of nAb against the specified virus variant. Antibody values assessed by means of live virus neutralising antibody assay that were reported as being below the lower limit of detection (LLOD; 8 for Ancestral SARS-CoV-2, Gamma, Delta, Omicron BA.1 and Omicron BA.5) were replaced by 0.5 times the LLOD.

[c] The 95% confidence intervals were calculated on the basis of the t-distribution of log-transformed values or difference in the log-transformed values for geometric mean titre and factor change in geometric mean titre, respectively, then back-transformed to the original scale

[d] GMFR: Fold change in the geometric mean titre respect to before booster antibody titres.

[e] 4× Seroconversion was defined as a change from below the LLOD to at least 4 times the LLOD, or an increase by a factor of at least four if the baseline value was greater than or equal to the LLOD; the comparison was with the baseline value. Percentages were based on the number of participants with nonmissing data at baseline and the corresponding time point; 95% confidence intervals were calculated with the use of the Wilson/Brown method.

[f] 10× Seroconversion was defined as a change from below the LLOD to at least 10 times the LLOQ, or an increase by a factor of at least ten if the baseline value was greater than or equal to the LLOQ; the comparison was with the baseline value. Percentages were based on the number of participants with nonmissing data at baseline and the corresponding time point; 95% confidence intervals were calculated with the use of the Wilson/Brown method.

**Table S3**

**Table S3. Comparison of the nAb response in male or female individuals after a booster dose of ARVAC-CG.**

| Variant | Dose | Sex | N[a] | nAb titre GMT (CI 95%)[b] d1 | | d14 | | P[d] | P[e] | 4× seroconversion[c] % | 95% CI | P[f] |
|---|---|---|---|---|---|---|---|---|---|---|---|---|
| Ancestral | 25 µg | M | 29 | **65·6** | (36·7-116·9) | **682·1** | (392·5-1185 | 0·0003 | 0·1 | **90·3** | 75·1 - 96·7 | 0·6197 |
| | | F | 31 | **70·0** | (42·5-115·3) | **1047** | (572·7-1915 | 0·0001 | | **86·2** | 69·4 - 94·5 | |
| | 50 µg | M | 8 | **26·9** | (7·3-99·2) | **861·1** | (262·1-2829 | 0·0004 | 0·8175 | **83·3** | 55·2 - 97·0 | 0·2235 |
| | | F | 12 | **25·4** | (10·7-60·5) | **724·1** | (240·3-2182 | 0·0002 | | **100·0** | 67·6 - 100·0 | |
| Gamma | 25 µg | M | 29 | **45·8** | (27·1-77·5) | **750·5** | (422·3-1334 | 0·0001 | 0·5764 | **90·3** | 75·1 - 96·7 | 0·9314 |
| | | F | 31 | **56·0** | (31·2-100·5) | **936·4** | (532·9-1645 | 0·0001 | | **89·7** | 73·6 - 96·4 | |
| | 50 µg | M | 8 | **19·0** | (5·4-67·6) | **664·0** | (238·4-1849 | 0·0007 | 0·7539 | **83·3** | 55·2 - 97·0 | 0·7982 |
| | | F | 12 | **28·5** | (11·4-71·3) | **812·7** | (315·8-2091) | 0·0001 | | **87·5** | 52·9 - 99·4 | |
| Delta | 25 µg | M | 29 | **32·8** | (19·7 - 54·6) | **403·1** | (238·7 - 680·8) | 0·0001 | 0·7538 | **93·1** | 78·0 - 98·8 | 0·0141 |
| | | F | 31 | **43·8** | (25·8 - 74·3) | **457·8** | (243·8 - 859·6) | 0·0001 | | **67·7** | 50·1 - 81·4 | |
| | 50 µg | M | 8 | **19·0** | (5·8 - 62·5) | **394·8** | (135·4 – 1151) | 0·0004 | 0·9657 | **87·5** | 52·9 - 99·4 | 0·7982 |
| | | F | 12 | **22·6** | (8·8 - 58·4) | **383·6** | (143·4 – 1026) | 0·0005 | | **83·3** | 55·2 - 97·0 | |
| Omicron BA.1 | 25 µg | M | 29 | **30·5** | (19·0-48·9) | **366·4** | (193·1-695·2) | 0·0001 | 0·5396 | **93·5** | 79·3 - 98·9 | 0·945 |
| | | F | 31 | **35·0** | (21·7-56·4) | **478·8** | (259·2-884·3) | 0·0001 | | **93·1** | 78·0 - 98·8 | |
| | 50 µg | M | 8 | **19·0** | (6·0-60·0) | **430·1** | (148·9-1245) | 0·022 | 0·1588 | **83·3** | 55·2 - 97·0 | 0·7982 |
| | | F | 12 | **26·9** | (9·0-80·6) | **966·5** | (461·9-2022) | 0·0001 | | **87·5** | 52·9 - 99·4 | |
| Omicron BA.5 | 25 µg | M | 29 | **8·8** | (6·3 - 12·3) | **62·5** | (39·3 - 99·5) | 0·0001 | 0·3692 | **79·3** | 61·6 - 90·2 | 0·8972 |
| | | F | 31 | **8·4** | (6·2 - 11·3) | **85·6** | (50·3 - 145·8) | 0·0001 | | **80·6** | 63·7 - 90·8 | |
| | 50 µg | M | 8 | **6·2** | (4·0 - 9·5) | **58·7** | (19·7 - 175·0) | 0·0004 | 0·2657 | **75·0** | 40·9 - 95·6 | 0·6481 |
| | | F | 12 | **8·0** | (4·1 - 15·8) | **128·0** | (46·6 - 351·9) | 0·0015 | | **83·3** | 55·2 - 97·0 | |
| **Variant** | **Dose** | **Sex** | **N** | **d1** | | **d28** | | **P[d]** | **P[b]** | **%** | **95% CI** | **P[c]** |
| Ancestral | 25 µg | M | 29 | **65·6** | (36·7-116·9) | **698·6** | (425·3-1147) | 0·0001 | 0·5254 | **76·7** | 59·1 - 88·2 | 0·1837 |
| | | F | 30 | **67·0** | (40·3-(111·1) | **601·9** | (344·1-1053) | 0·003 | | **89·7** | 73·6 - 96·4 | |
| | 50 µg | M | 7 | **26·3** | (5·5-124·6) | **512·0** | (206·8-1268) | 0·0034 | 0·5013 | **81·8** | 52·3 - 96·8 | 0·8288 |
| | | F | 11 | **28·2** | (11·2-71·29) | **329·4** | (120·6-899·5) | 0·002 | | **85·7** | 48·7 - 99·3 | |
| Gamma | 25 µg | M | 29 | **45·8** | (27·1-77·46) | **443·6** | (269-731·5) | 0·0001 | 0·936 | **70·0** | 52·1 - 83·3 | 0·2495 |
| | | F | 30 | **54·4** | (29·8-99·49) | **456·1** | (276·7-751·9) | 0·0001 | | **82·8** | 65·5 - 92·4 | |
| | 50 µg | M | 7 | **17·7** | (3·9-79·18) | **344·6** | (130·7-908·2) | 0·003 | 0·8575 | **81·8** | 52·3 - 96·8 | 0·8288 |
| | | F | 11 | **32·0** | (12·1-84·99) | **309·3** | (127·5-749·9) | 0·0033 | | **85·7** | 48·7 - 99·3 | |
| Omicron | 25 µg | M | 29 | **68·8** | (39·6-119·4) | **288·5** | (147·3-565·2) | 0·0002 | 0·8776 | **51·5** | 35·2 - 67·5 | 0·908 |
| | | F | 30 | **67·0** | (40·3-111·4) | **308·0** | (178·1-532·6) | 0·0001 | | **50·0** | 32·1 - 67·9 | |
| | 50 µg | M | 7 | **23·8** | (5·7-99·02) | **231·9** | (97·9-549·2) | 0·0022 | 0·2993 | **81·8** | 52·3 - 96·8 | 0·6052 |
| | | F | 11 | **30·5** | (9·2-98·41) | **423·8** | (179·2-1002) | 0·002 | | **71·4** | 35·9 - 94·9 | |

[a] Number of participants with nonmissing data at the time point (or at baseline).

[b] GMT: Geometric Mean titre of nAb against the specified virus variant. Antibody values assessed by means of live virus neutralising antibody assay that were reported as being below the lower limit of detection (LLOD; 8 for Ancestral SARS-CoV-2, Gamma, Delta, Omicron BA.1 and Omicron BA.5) were replaced by 0.5 times the LLOD. The 95% confidence intervals were calculated on the basis of the t-distribution of log-transformed values or difference in the log-transformed values for geometric mean titre and factor change in geometric mean titre, respectively, then back-transformed to the original scale.

[c] 4× Seroconversion was defined as a change from below the LLOD to at least 4 times the LLOD, or an increase by a factor of at least four if the baseline value was greater than or equal to the LLOD; the comparison was with the pre-vaccination baseline value. Percentages were based on the number of participants with nonmissing data at baseline and the corresponding time point; 95% confidence intervals were calculated with the use of the Wilson/Brown method.

[d] Exact P Value. d1 vs. d14 or d28, respectively. Wilcoxon pair-matched test.

[e] Exact P value. D14 or d28 M vs d12 or d28 F. Mann Whitney test t.

[f] Exact P value. M vs F. Fisher's exact test.

### Table S4

| Table S4: Neutralising antibody and seroconversion Analysis against Ancestral SARS-CoV-2 (Wuhan), Gamma, and Omicron BA.1 after 28 days of administration of 25 or 50 µg of ARVAC-CG as Booster Dose in Participants. | | | | | | |
|---|---|---|---|---|---|---|
| **SARS-CoV-2 variant** | **Ancestral** | | **Gamma** | | **Omicron BA·1** | |
| **ARVAC-CG Dose** | **25 µg** | **50 µg** | **25 µg** | **50 µg** | **25 µg** | **50 µg** |
| No. of participants evaluated[a] | (N=58) | (N=18) | (N=58) | (N=18) | (N=58) | (N=18) |
| **Before booster** | | | | | | |
| GMT[b] | **69·6** | **27·4** | **52·2** | **25.4** | **33.6** | **23.5** |
| (95% CI)[c] | (48·25-100·4) | (13·3-56·6) | (35·5-76·86) | (12.0-53.6) | (24.16-46.64) | (10.5-52.5) |
| **Day 28** | | | | | | |
| GMT[b] | **658·1** | **391·0** | **443·6** | **322.5** | **299.0** | **335.2** |
| (95% CI)[c] | (455·1-951·5) | (204·3-748·3) | (313·3-628·1) | (178.6-582.5) | (195.2-458.1) | (187.5-599.2) |
| GMFR[d] | **9·5** | **14·3** | **8·5** | **12.7** | **8.9** | **14.3** |
| (95% CI)[c] | (6·5-13·1) | (5·7-35·4) | (5·8-12·4) | (5.1-31.5) | (6.1-13.0) | (5.9-34.3) |
| Mann Whitney test. P | 0·1867 (ns) | | 0·2179 (ns) | | 0·1848 (ns) | |
| **4× Seroconversion at day 28[e]** | | | | | | |
| **Percentage of participants** | **82·8** | **83·3** | **75·9** | **83.3** | **75.9** | **83.3** |
| **(95% CI)** | (71·1-90·4) | (60·8-94·2) | (63·5-85·0) | (60.8-94.2) | (63.5-85.0) | (60.8-94.2) |
| Fisher's exact test, P | >0·9999 (ns) | | 0·7474 (ns) | | 0·7474 (ns) | |
| Chi-square test, P | 0·9549 (ns) | | 0·5064 (ns) | | 0·5064 (ns) | |
| **10× Seroconversion at day 28[f]** | | | | | | |
| **Percentage of participants** | **39·7** | **72·2** | **43·1** | **61.1** | **37.9** | **61.1** |
| **(95% CI)** | (28·1-52·5) | (49·1-87·5) | (31·2-55·9) | (38.6-79.7) | (26.6-50.8) | (38.6-79.7) |
| Fisher's exact test, P | 0·0289 (*) | | 0·2798 (ns) | | 0·1058 (ns) | |
| Chi-square test, P | 0·0156 (*) | | 0·1813 (ns) | | 0·083 (ns) | |

[a] Shown is the number of participants with nonmissing data at the time point (or at baseline).

[b] Antibody values assessed by means of live virus neutralising antibody assay that were reported as being below the lower limit of detection (LLOD; 8 for Ancestral SARS-CoV-2, Gamma, Delta, Omicron BA.1 and Omicron BA.5) were replaced by 0.5 times the LLOD.

[c] The 95% confidence intervals were calculated on the basis of the t-distribution of log-transformed values or difference in the log-transformed values for geometric mean titre and factor change in geometric mean titre (GMT), respectively, then back-transformed to the original scale.

[d] GMFR: Fold change in the GMT titre respect to baseline antibody titres.

[e] 4× Seroconversion was defined as a change from below the LLOD to at least 4 times the LLOD, or an increase by a factor of at least four if the baseline value was greater than or equal to the LLOD; the comparison was with the baseline value. Percentages were based on the number of participants with no missing data at baseline and the corresponding time point; 95% confidence intervals were calculated with the use of the Wilson/Brown method.

[f] 10× Seroconversion was defined as a change from below the LLOD to at least 10 times the LLOD, or an increase by a factor of at least ten if the baseline value was greater than or equal to the LLOD; the comparison was with the baseline value. Percentages were based on the number of participants with no missing data at baseline and the corresponding time point; 95% confidence intervals were calculated with the use of the Wilson/Brown method.

### Supplementary Methods

#### Determination of SARS-CoV-2 Nucleoprotein (N)-specific antibody levels in serum.

Nucleoprotein (N)-specific antibody responses (IgG) were evaluated by indirect ELISA. Recombinant N protein (0.25 µg/well) was used to coat plates and HRP conjugated anti-human IgG was used to detect Ab. Results were read at 450 nm to collect endpoint ELISA data. End-point cut-off values for serum titre determination were calculated as the mean specific optical density (OD) plus 3 standard deviations (SD) from pre-pandemic sera of healthy donors diluted 1:100 in assay diluent. Titres were established as the reciprocal of the last dilution yielding an OD higher than the cut-off.
